# CLOT STIFFNESS MEASURED BY SEER SONORHEOMETRY AS A MARKER OF POOR PROGNOSIS IN HOSPITALIZED COVID-19 PATIENTS

**DOI:** 10.1101/2021.07.13.21260256

**Authors:** Francisco José López-Jaime, Sandra Martín-Téllez, Alberto Doblas-Márquez, Ignacio Márquez-Gómez, José María Reguera-Iglesias, Manuel Isidro Muñoz-Pérez, Ihosvany Fernández-Bello, Adrián Montaño

## Abstract

**Background:** High incidence of life-threatening thrombotic complications is observed in severely ill COVID-19 patients. D-dimer may help evaluate disease severity and predict outcomes at hospital admission. However, its non-specificity and long analysis times strongly constrain its clinical value. Viscoelastic tests (VET) are widely available rapid point-of-care devices that have been shown to detect a hypercoagulable state (increased clot stiffness and fibrinolysis shutdown) as major contributors of the thrombotic complication in COVID-19. Nevertheless, based on the data obtained so far, definitive conclusions have not been drawn.

**Objectives:** We aim to evaluate the association between VET parameters, standard coagulation tests and inflammation markers assessed in COVID-19 patients at hospital admission with disease severity and outcomes.

**Patients/Methods:** A total of 69 COVID-19 patients requiring hospitalization were included in the study. The pro-inflammatory and pro-thrombotic state was analyzed by a panel of inflammation markers (IL-6, CRP, LDH, ferritin), routine coagulation tests (PT, aPTT, platelet count, fibrinogen, D-dimer) and a SEER sonorheometry VET profile (Quantra^®^ System).

**Results:** Inflammatory markers IL-6, CRP, LDH and ferritin were elevated in a high percentage of patients (73.6%, 89.2%, 57.1% and 52.4%), as were coagulation-related parameters such as fibrinogen (81.4%) and D-dimer levels (66.2%). Quantra^®^ analysis revealed increased clot stiffness (CS) in 34.8%, particularly due to increased fibrinogen contribution (FCS) in 63.7%. Increased clot stability to lysis (CSL) was observed in 32.4%. Age > 65 years, elevated values of fibrinogen, D-dimer, LDH, increased clot stiffness and resistance to clot lysis were significantly associated with worsening disease. The Quantra^®^ FCS parameter showed a particularly high prognostic value in distinguishing patients with severe symptomatology.

**Conclusion:** The global study of hemostasis by the whole blood point-of-care Quantra^®^ VET system may be a powerful tool for identifying poor prognosis in COVID-19 patients at hospital admission. In particular, FCS measured by Quantra^®^ could be established as a plausible prognostic marker to aid the clinical management of COVID-19 patients.

## 1. INTRODUCTION

At the moment of writing this article, coronavirus disease 2019 (COVID-19) caused by severe acute respiratory syndrome coronavirus 2 (SARS-CoV-2) has already killed more than 3 million people worldwide [1]. The development of an extreme inflammatory response and prothrombotic state is associated with poor prognosis and remains the major cause of the high morbidity and mortality rate associated with this condition [2,3].

Previous studies on the hemostatic status of COVID-19 patients identified several parameters such as fibrinogen and D-dimer that could be established as markers of thrombotic risk and helped in the clinical management of patients [3–5]. Elevated D-dimer levels (>1000 ng/mL) have also been proposed as an indicator of risk for pulmonary embolism [6]. However, these parameters have drawbacks that affect their reliability and clinical use to predict outcomes in COVID-19. D-dimer in particular has important limitations that raises questions on its true prognostic value for thrombosis in COVID-19. Although highly elevated levels of D-dimer in COVID-19 have been attributed to acute lung injury with intra-alveolar fibrin deposition and subsequent breakdown of this extravascular fibrin, D-dimer values may be elevated for a variety of reasons and therefore increased values are not only indicative of increased thrombotic risk [7–10]. In addition, D-dimer requires a long time of analysis which together weaken its clinical value and reveals the need to identify new biomarkers that provide a rapid and effective assessment of thrombotic risk in COVID-19.

Viscoelastic tests (VET) are recognized powerful tools for the rapid identification and characterization of hypercoagulable state in infectious diseases and several groups have already evaluated the use of VET in COVID-19 patients [11,12]. In general, the available data indicate a high frequency of increased clot stiffness and fibrinolysis shutdown but with weak evidence on the association of this hypercoagulable profile with disease severity. Larger studies with larger numbers of patients are therefore needed to confirm these important findings.

Sonic Estimation of Elasticity by Resonance (SEER) sonorheometry (Quantra® system) is a novel ultrasound-based VET technology [13]. With this system, a hypercoagulable profile has been reported in hospitalized COVID-19 patients [14,15]. The objective of this study is to investigate whether the Quantra parameters, as well as a panel of inflammation markers and routine coagulation tests, are associated with disease severity and outcomes of hospitalized COVID-19 patients.

## 2. MATERIALS AND METHODS

### 2.1 Patients and diagnosis criteria

A total of 69 patients, positive for SARS-CoV-2 by PCR and hospitalized at the Regional University Hospital of Málaga were included in the study. All patients were over 18 years of age and none had any other hemostasis disorders. Immunocompromised patients, such as oncology patients on chemotherapy or those positive for HIV were also excluded from the study.

Patients were classified according to disease severity (critical, severe, and non-severe) based on the World Health Organization (WHO) criteria [16]. Briefly, critical COVID-19 patients were those who presented sepsis, septic shock or other conditions that required provision of life-sustaining therapies (mechanical ventilation or vasopressor therapy); severe were those with oxygen saturation <90% on room air or respiratory rate >30 breaths/min or with signs of severe respiratory distress including the need to use accessory muscles and the inability to formulate full sentences; and finally, the absence of any critical or severe COVID-19 criteria was considered as non-severe disease. Patients were also classified according to the presence or absence of adverse events, including the need for mechanical ventilation, development of encephalitis, thrombotic events, admission to ICU (intensive care unit) or death. The study was approved by the local ethical committee. Written informed consent was obtained from patients or legal guardians before inclusion.

### 2.2 Inflammatory and procoagulant status

Citrated, EDTA and non-anticoagulated blood samples were drawn on hospital admission and after confirmation of SARS-CoV-2 infection. Quantra^®^ Hemostasis Analyzer (HemoSonics, Charlottesville, VA, USA) was used to evaluate the global viscoelastic hemostatic status of the patients. On this system, QStat^®^ and QPlus^®^ cartridges were used to determine the following parameters: Clot Time (CT), Heparinase Clot Time (CTH), Clot Time Ratio (CTR: CT/CTH), Clot Stiffness (CS), Fibrinogen Contribution to Clot Stiffness (FCS), Platelet Contribution to Clot Stiffness (PCS), Clot Stability to Lysis (CSL). Details of Quantra QPlus and QStat cartridges have been described previously [17,18]. Complete blood count, prothrombin time (PT), international normalized ratio (INR), activated partial thromboplastin time (aPTT), D-dimer, fibrinogen, IL-6, ferritin, lactate dehydrogenase (LDH) and C-reactive protein (CRP) were also determined.

### 2.3 Statistical analysis

The statistical software “SPSS statistics” (IBM) was used for the analysis. Comparison for qualitative (Chi-square test or Fisher’s Exact Test) and quantitative differences (Mann-Whitney U test) were applied. Relationship between parameters at admission and disease severity or adverse events was assessed by ROC curve analysis.

## 3. RESULTS

### 3.1 Demographic and clinical characteristics of the patients

Table 1 and supplementary file 1 show demographic and clinical characteristics of the patient cohort: 44.9% of patients developed non-severe disease, 39.1% severe disease and 16% critical disease. Compared to non-severe patients, severe/critical patients were older, often female and had more diabetes and adverse events. A total of 26.1% of the patients had an adverse event during hospital admission. Except for encephalitis, all adverse events occurred in the severe/critical group.

**Table 1.**
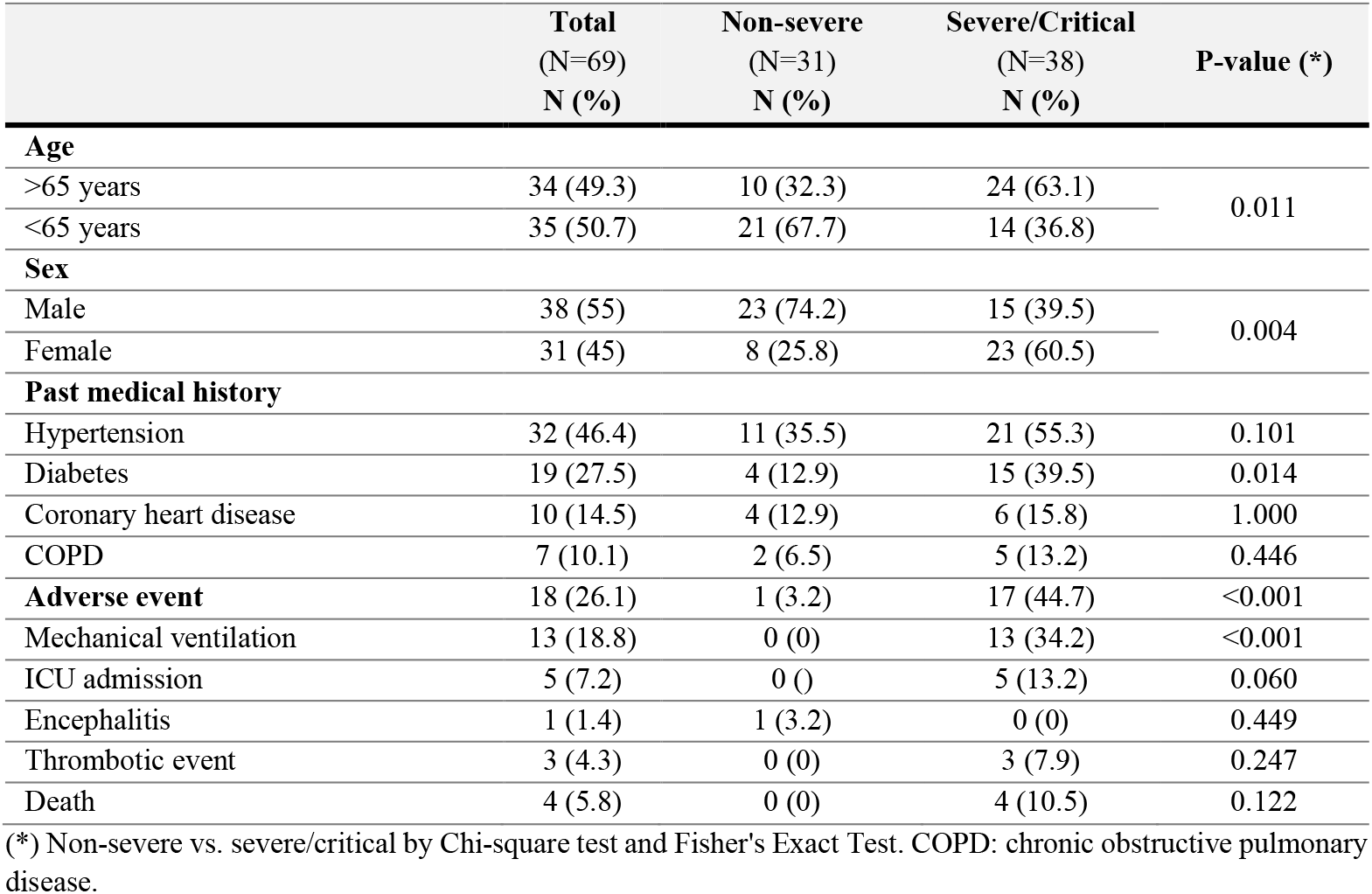
Demographic and clinical characteristics of COVID-19 patients with non-severe disease versus severe/critical disease.

|  | Total<br>(N=69)<br>N (%) | Non-severe<br>(N=31)<br>N (%) | Severe/Critical<br>(N=38)<br>N (%) | P-value (*) |
| --- | --- | --- | --- | --- |
| **Age** |  |  |  |  |
| >65 years | 34 (49.3) | 10 (32.3) | 24 (63.1) | 0.011 |
| <65 years | 35 (50.7) | 21 (67.7) | 14 (36.8) |  |
| **Sex** |  |  |  |  |
| Male | 38 (55) | 23 (74.2) | 15 (39.5) | 0.004 |
| Female | 31 (45) | 8 (25.8) | 23 (60.5) |  |
| **Past medical history** |  |  |  |  |
| Hypertension | 32 (46.4) | 11 (35.5) | 21 (55.3) | 0.101 |
| Diabetes | 19 (27.5) | 4 (12.9) | 15 (39.5) | 0.014 |
| Coronary heart disease | 10 (14.5) | 4 (12.9) | 6 (15.8) | 1.000 |
| COPD | 7 (10.1) | 2 (6.5) | 5 (13.2) | 0.446 |
| **Adverse event** | 18 (26.1) | 1 (3.2) | 17 (44.7) | <0.001 |
| Mechanical ventilation | 13 (18.8) | 0 (0) | 13 (34.2) | <0.001 |
| ICU admission | 5 (7.2) | 0 (0) | 5 (13.2) | 0.060 |
| Encephalitis | 1 (1.4) | 1 (3.2) | 0 (0) | 0.449 |
| Thrombotic event | 3 (4.3) | 0 (0) | 3 (7.9) | 0.247 |
| Death | 4 (5.8) | 0 (0) | 4 (10.5) | 0.122 |

(\*) Non-severe vs. severe/critical by Chi-square test and Fisher's Exact Test. COPD: chronic obstructive pulmonary disease.

### 3.2 Increased clot stiffness in patients with poor prognosis

As shown in Table 2, patients with adverse events showed significantly higher values of CRP, LDH, fibrinogen, D-dimer and the Quantra clot stiffness parameter FCS.

**Table 2.**
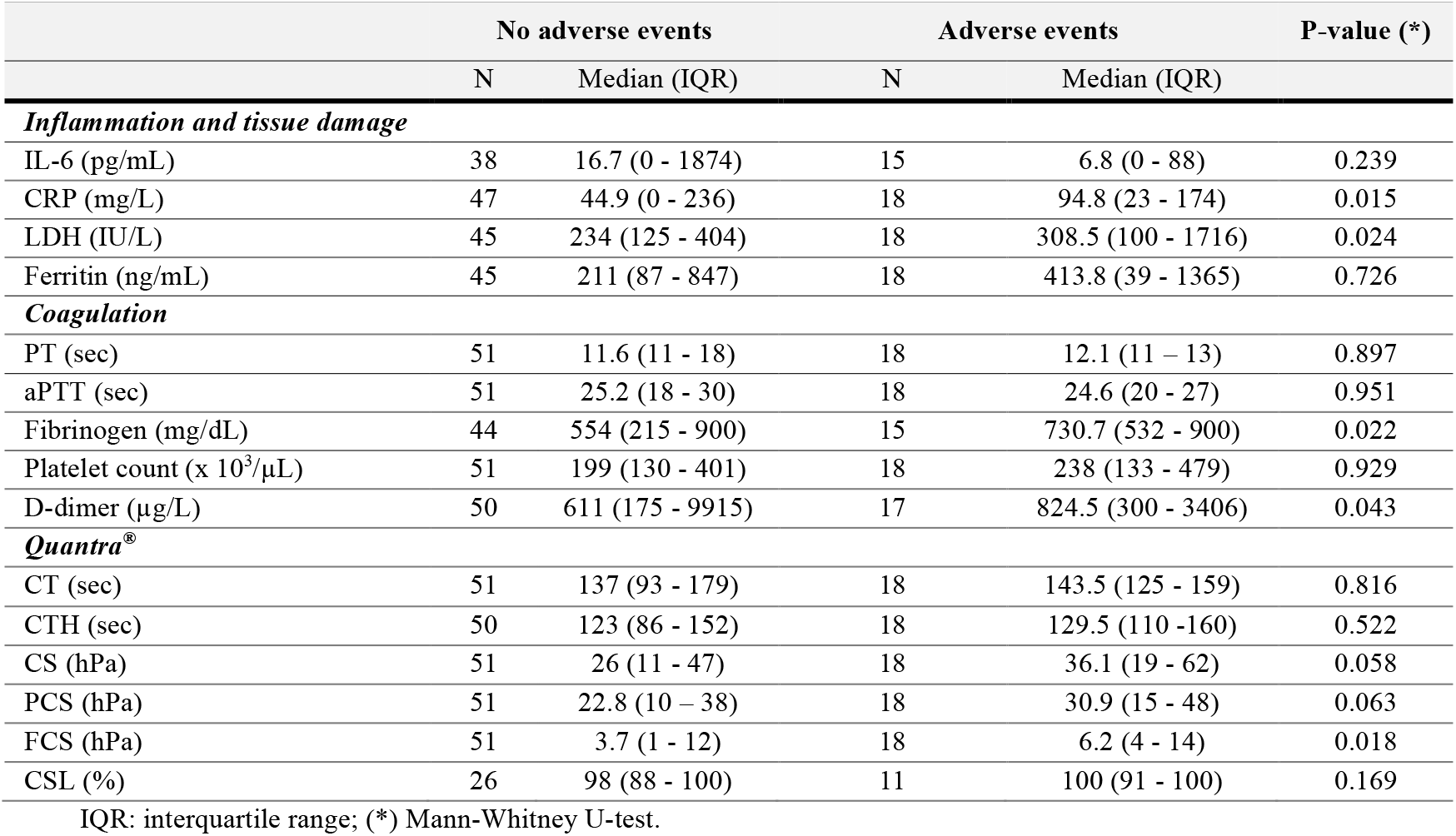
Median of inflammation and tissue damage markers, coagulation and Quantra^®^ parameters in patients with and without adverse events.

Table 3 shows the proportion of elevated levels of all assessed parameters, defined as the proportion above the upper limit of normal (ULN), in the total cohort as well as in patients who did not experience an adverse event and those who did. Inflammation and tissue damage markers were elevated in at least half of the patients: IL-6 in 73.6%, CRP in 89.2%, LDH in 57.1% and ferritin in 52.4%. Regarding coagulation parameters, shortened PT and aPTT clot times, i.e., below the lower limit of normal (LLN), were only observed in 0% and 14.5%, respectively. An elevated platelet count was only present in 8.7% of all patients. In contrast, fibrinogen and D-dimer were elevated in 81.4% and 66.2%, respectively. On the Quantra system, shortened CT and CTH clot times (below LLN) were only observed in 8.7% and 14.7%, respectively. Total clot stiffness (CS) was elevated in 34.8%, with an elevated PCS and FCS in 27.6% and 63.7%, respectively. A CSL value of 100% (suggesting absence of fibrinolysis) was observed in 32.4%.

**Table 3.** Markers of inflammation and tissue damage, coagulation and Quantra parameters: proportion above upper limit of normal (ULN) or below the lower limit of normal (LLN) in COVID-19 patients with no adverse events versus those with adverse events. In the case of PT, aPTT, CT and CTH, the percentage of patients with times below the lower limit of normal values was considered.

| Test | ULN/<br>BLN | N/total (%) | No adverse<br>events<br>N/total (%) | Adverse<br>events<br>N/total (%) | P-value<br>(*) |
| --- | --- | --- | --- | --- | --- |
| **Inflammation and tissue damage** |  |  |  |  |  |
| IL-6 (pg/mL) | 4.4 | 39/53 (73.6) | 28/38 (73.7) | 11/15 (73.3) | 1.000 |
| CRP (mg/L) | 5.0 | 58/65 (89.2) | 41/47 (87.2) | 17/18 (94.4) | 0.663 |
| LDH (IU/L) | 246 | 36/63 (57.1) | 21/45 (46.7) | 15/18 (83.3) | 0.008 |
| Ferritin (ng/mL) | 388 | 33/63 (52.4) | 22/45 (48.9) | 11/18 (61.1) | 0.380 |
| **Coagulation** |  |  |  |  |  |
| PT (sec) | 10.0 | 0/69 (0) | 0/51 (0) | 0/18 (0) | 1.000 |
| aPTT (sec) | 20.6 | 10/69 (14.5) | 8/51 (15.7) | 2/18 (11.1) | 1.000 |
| Platelet count ($\times 10^3/\mu\text{L}$) | 450 | 6/69 (8.7) | 5/51 (9.8) | 1/18 (5.6) | 1.000 |
| Fibrinogen (mg/dL) | 400 | 48/59 (81.4) | 34/44 (77.3) | 14/15 (93.3) | 0.259 |
| D-dimer ($\mu\text{g/L}$) | 500 | 45/68 (66.2) | 29/50 (58) | 16/18 (88.9) | 0.018 |
| **Quantra®** |  |  |  |  |  |
| CT (sec) | 113 | 6/69 (8.7) | 3/51 (5.9) | 2/18 (11.1) | 0.600 |
| CTH (sec) | 109 | 10/68 (14.7) | 7/50 (14) | 3/18 (16.7) | 0.717 |
| CS (hPa) | 33.2 | 24/69 (34.8) | 14/51 (27.4) | 10/18 (55.6) | 0.031 |
| PCS (hPa) | 29.8 | 19/69 (27.6) | 11/51 (21.6) | 8/18 (44.4) | 0.074 |
| FCS (hPa) | 3.7 | 44/69 (63.7) | 28/51 (54.9) | 16/18 (88.9) | 0.008 |
| CSL (%) | 100 | 12/37 (32.4) | 5/26 (19.2) | 7/11 (63.6) | 0.018 |

(\*) Non-severe vs. severe/critical by Chi-square test and Fisher's Exact Test.

Only, LDH, D-dimer, the Quantra clot stiffness parameters (CS and FCS) and resistance to lysis (CSL) showed a significantly higher proportion of elevated values in COVID-19 patients with no adverse events compared with those with adverse events. Patients with elevated FCS values together with elevated D-dimer values were significantly associated with the presence of an adverse event (P<0.001). In predicting the severity of disease, the area under the curve (AUC) by ROC analysis was 0.720 with a 95% confidence interval of [0.546 – 0.895] (*P*=0.038) for FCS; 0.581 [0.379 – 0.783] (*P*=0.443) for LDH; 0.635 [0.439 – 0.831] (*P*=0.203) for D-dimer; 0.673 [0.485 – 0.860] (*P*=0.104) for CS and 0.538 [0.336 – 0.739] (*P*=0.722) for CSL. In predicting the manifestation of an adverse event, the AUC was 0.715 [0.541 – 0.888] (*P*=0.047) for FCS; 0.645 [0.440 – 0.849] (*P*=0.181) for LDH; 0.562 [0.361 – 0.763] (*P*=0.567) for D-dimer; 0.657 [0.451 – 0.863] (*P*=0.147) for CS and 0.643 [0.422 – 0.863] (*P*=0.188) for CSL. Thus, only FCS showed the highest significant AUC compared to the other variables such as D-dimer, as shown in figure 1 (*P*=0.003).

**Figure 1.**
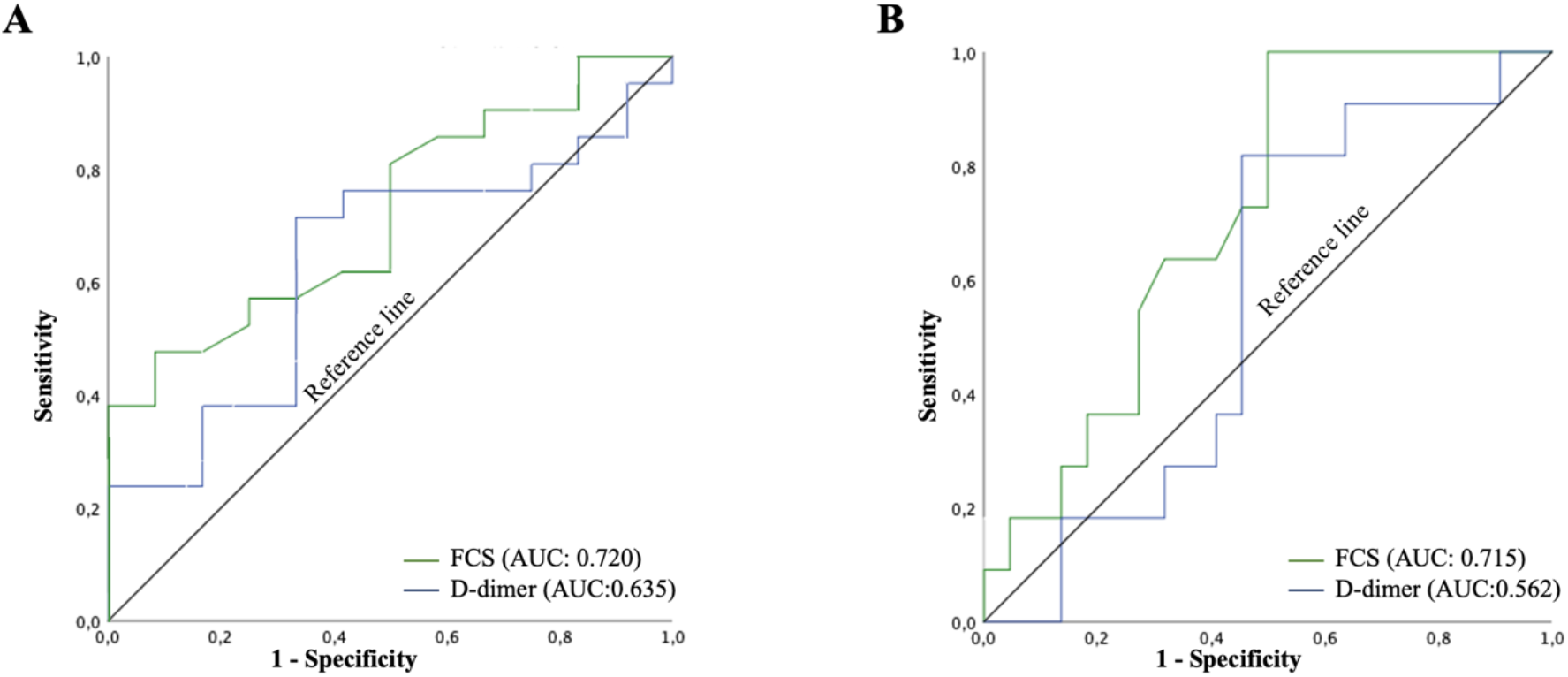
ROC analysis. **A)** ROC curve of FCS and D-dimer at admission in predicting severity of disease. **B)** ROC curve of FCS and D-dimer at admission in predicting the manifestation of an adverse event.

### 3.3 Close interaction between procoagulant status and inflammation in COVID-19 patients

FCS values showed a strong correlation with plasma fibrinogen (r=0.850; *P*<0.001). In turn, both parameters showed a moderate to strong correlation with CRP values (0.699; *P*<0.001 for FCS and 0.803; *P*<0.011 for plasma fibrinogen). Elevated FCS and plasma fibrinogen values were significantly associated with elevated CRP values (*P*=0.008 and *P*<0.001, respectively) and IL-6 values (*P*=0.028 and *P*<0.001, respectively). On the other hand, PCS correlated strongly with platelet count (r=0.752; *P*<0.001) (Figure 2).

**Figure 2.**
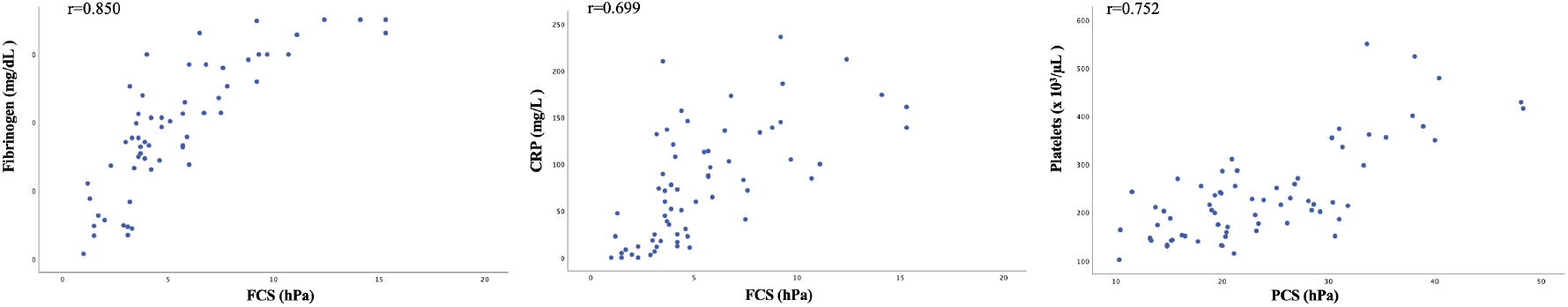
Scatter plots and Spearman correlation. FCS vs. fibrinogen (left) and CRP (middle); PCS vs. platelet count (right).

## 4. DISCUSSION

We present the results of a pilot study in which we evaluated the use of global viscoelastic hemostatic analysis by sonorheometry (Quantra system) in COVID-19 patients on admission and the association of the Quantra parameters with disease severity and outcomes. Whole blood samples from 69 COVID-19 patients were analyzed using standard laboratory tests for coagulation, inflammatory and tissue damage markers and the Quantra Hemostasis Analyzer. Global hemostatic analysis by the Quantra revealed increased clot stiffness in a high number of COVID-19 patients, especially in those who experienced an adverse event during admission, such as the need for mechanical ventilation, development of encephalitis, thrombotic event, ICU admission or death. Particularly the FCS value was shown to indicate poor prognosis and could be of great use in the clinical management of these patients, both in triage and in the development of new therapeutic approaches.

Of the total cohort, 55.1% were classified as having severe/critical disease according to WHO criteria [16]. As expected, all patients who experienced an adverse event belonged to this group, except for one patient in the non-severe disease group who developed encephalitis. The most occurring event was the presence of ARDS observed in 18.8% of patients, who required mechanical ventilation. Consistent with previous studies, we observed that an age > 65 years was indicative of a worse prognosis [19,20]. Thus, patients with advanced age were more frequently observed in the severe/critical disease group. Other comorbidities such as diabetes were also associated with a worse prognosis as previously described [21,22].

We observed in our series a high proportion of patients with IL-6 and CRP values above the ULN. In support of our finding, these markers have been shown to be elevated in COVID-19 patients, especially in those with ARDS [5]. In fact, the group of Herold. et al. demonstrated the value of these parameters as predictors of the need for mechanical ventilation. Elevated fibrinogen levels were also associated with elevated Il-6 values, thus establishing a clear interaction between coagulation and inflammation, as reported in the study by Ranucci et al [14]. This is further evidence in support of the concept of immunothrombosis as a pathogenetic mechanism in COVID-19 [20]. A high percentage of patients in our cohort also showed LDH values above ULN, especially in those associated with worse prognosis, consistent with lung damage due to pneumonia caused by SARS-CoV-2 infection and which has been associated with increased patient mortality [23].

Regarding coagulation parameters, COVID-19 patients showed elevated levels of fibrinogen and D-dimer, the latter being mostly observed in patients who experienced an adverse event during admission. These results are also in agreement with those reported in the literature, which propose these factors as probably responsible for the thrombotic events that have been observed in COVID-19 patients [3,4].

VET analysis has also been proposed as a powerful tool for the study of the global hemostatic status of COVID-19 patients [24]. Especially, these studies have focused on the analysis of patients with poor prognosis such as those admitted to the ICU or with ARDS, revealing that VET analysis could be established as a promising tool to predict thrombotic complications and thus optimize the diagnosis and treatment of COVID-19 patients [25]. Our results showed that a high percentage of patients had altered parameters of clot stiffness, mainly due to the contribution of fibrinogen. Alterations in these parameters were significantly more prevalent in patients with an adverse event during admission. These results support previous studies suggesting that the hypercoagulability of COVID-19 patients is largely attributable to the contribution of fibrin and platelets to clot stability and stiffness [14].

As expected, platelet count and fibrinogen levels correlated positively with PCS and FCS values, respectively. However, despite the high correlation between platelet count and PCS, an elevated platelet count was observed in only 8.7% of patients, compared to 27.6% of patients with an elevated PCS. This result supports the evidence shown by Baryshnikova et al., that PCS is not only influenced by platelet count, but also by platelet functionality [26]. Furthermore, these findings highlight that platelet hyperreactivity plays a key role in the pathogenicity of severely ill COVID-19 patients, as previous studies have shown [27]. On the Quantra QStat, based on response in normal healthy individuals, the reference range is 93% to 100%. In our cohort tested for CSL (n=37; 11 with adverse events), only 4 patients (1 with adverse events) showed a value below the lower limit of normal, suggesting hyperfibrinolysis. Although for the QStat CSL parameter a cut-off for fibrinolysis shutdown has not been determined, it is interesting to observe that the proportion with the maximum CSL value of 100% was significantly higher in the patients with an adverse event (63.6% vs. 19.2%; *P*=0.018). This finding suggests the presence of fibrinolysis resistance in support of previous findings that attributed the hypercoagulability of COVID-19 patients to this phenomenon [28–30].

Many of these parameters, both inflammation and coagulation, have been proposed as promising prognostic markers to help us predict the risk of COVID-19 patients. In particular, many studies have focused on D-dimer as an indicator of poor prognosis [20,31]. However, the prognostic value of D-dimer has several limitations such as long analysis times, highlighting the need to identify other powerful markers. On the other hand, increased clot stiffness has been observed especially in patients with ARDS or ICU admission, revealing a possible association between these parameters and a worse prognosis of the disease [32]. Nevertheless, the prognostic value of clot stiffness measured by VET techniques has not yet been extensively studied. Our results demonstrate that clot stiffness parameters measured by Quantra^®^ had a high prognostic value according to ROC analysis. In particular, fibrinogen contribution to clot stiffness (FCS) had the highest AUC compared to other parameters strongly associated with worse prognosis such as age or D-dimer. These findings highlight the potential use of this marker as predictor of poor prognosis in COVID-19 patients.

One of the limitations of our study is, however, that patients were analyzed only at the time of diagnosis. Previous studies have shown that the predictive value of D-dimer on the risk of death is much higher when measured just before the clinical outcome compared to the time of diagnosis, suggesting the importance of dynamically monitoring D-dimer levels to detect thrombotic complications and thus reduce the mortality rate of patients [20]. The results of our study, which found that FCS has a higher predictive value for an adverse event than D-dimer, indicate that FCS may also be an important parameter to monitor, especially in COVID-19 patients with elevated FCS at hospital admission. In this study we also observed that both elevated FCS and D-dimer were significantly more strongly associated with patients with a worse prognosis, suggesting the joint use of both parameters as a predictor of risk.

In summary, we can conclude from this study that the use of the sonorheometry VET technique allows a global analysis of the hemostatic status of COVID-19 patients in a rapid and efficient way, including clot stiffness parameters (CS, PCS, FCS, CSL) that are not routinely measured by standard laboratory tests. The implementation of this technique would allow for a more accurate diagnosis of patients on admission, helping in patient triage. By using this technique, we were able to detect that COVID-19 patients with higher clot stiffness showed a worse prognosis, especially due to the higher contribution of fibrinogen. Although further studies are needed to confirm these findings, these results show that the detection of FCS at admission could be of particular interest in the clinical management of COVID-19 patients and could also contribute to the development of new therapeutic strategies thanks to the numerous anticoagulant agents currently available.

## Data Availability

No data in external datasets

## AUTHOR CONTRIBUTIONS

Conceptualization, F.J.L.-J, M.I.M.-P and I.F.-B; Methodology, F.J.L.-J, I.M.-G, I.F.-B and A.M; Software, F.J.L.-J and A.M; Validation, S.M.-T, I.F.-B and A.M; Formal analysis, F.J.L.-J, I.F.-B and A.M; Investigation, F.J.L.-J, S.M.-T, A.D.-M, J.M.R.-I and A.M; Data curation, A.D.-M, I.F.-B and A.M; Writing – original draft preparation, F.J.L.-J, I.F.-B and A.M, Writing – review and editing, all authors; Visualization, all authors; Supervision; F.J.L.-J and I.F.-B; Project administration, F.J.L.-J; Funding acquisition, F.J.L.-J; All authors have read and agreed to the published version of the manuscript.

## ACKNOWLEDGEMENTS

The authors would like to thank to Beatriz Gago Caballero, Jessica Aguilera Rodríguez, Margarita López Quintero and Cristina Montes López from the Hospital Regional Universitario of Málaga, Spain, for their technical assistance. We would also like to thank all the nurses in the infectious diseases area of this hospital for their technical assistance. The study has been carried out in cooperation with Hemosonics but the company had no influence on decision to publish.

